# Population-Scale Integration of Spatial Omics Networks for Clinical Prediction and Biological Discovery by SPIN

**DOI:** 10.1101/2025.08.09.25333297

**Authors:** Tong Liu, Euiseong Ko, Yiwen Yang, Haowen Wu, Thatchayut Unjitwattana, Suyuan Wang, Jian Kang, Yijun Li, Lana X Garmire

## Abstract

Spatial omics enables the integration of high-dimensional molecular organization with clinical outcomes, yet incorporating spatial single-cell information into predictive models at the population scale remains challenging. Here, we introduce SPIN (Spatial Predictive Integration Network), which integrates subject-specific, spatially informed co-expression or co-abundance networks for population-scale clinical prediction. Current implementation of SPIN includes three key modules: network construction, molecular-to-clinical prediction by Bayesian scalar-on-network regression with manifold learning (BSNMani), and visualization & interpretation module. In the SEA-AD MERFISH transcriptomics cohort, SPIN framework achieved an accuracy of 0.81 for dementia-status prediction and revealed gene–gene co-expression subnetworks with clear biological relevance, such as glutamatergic synapses- and neurogenesis-related subnetworks. SPIN also achieved robust survival prediction in a breast cancer single proteomics cohort (C-index=0.78), identifying two survival-associated spatial proteomic subnetworks. SPIN can further use cell-type-specific spatial omics data to enhance the granularity. In summary, SPIN is a valuable tool that uses high-dimensional spatial omics data for clinical outcome prediction at the population scale across diverse disease settings, revealing biological insights while maintaining interpretation. SPIN package is available at: https://github.com/lanagarmire/SPIN

## Introduction

Recent advances in spatial transcriptomics (ST) have enabled the measurement of gene expression at near-single-cell resolution while preserving the spatial context within tissues. These technologies provide unprecedented opportunities to investigate not only the molecular states of individual cells but also their spatial organization, interactions, and microenvironments [1,2]. As a result, ST has transformed our ability to characterize tissue architecture and cellular heterogeneity in healthy and diseased tissues. Many computational methods have been developed to characterize gene expression variation across tissue space [3–5], identify spatial domains [6–8], and construct cell-cell interaction profiles in tissue niches [9,10]. Together, these approaches have deepened our understanding of the spatial organization of gene activity within individual tissue samples.

One promising computational avenue for extracting interpretable, high-level structure from gene expression data is through co-expression networks. These methods group genes into functional modules, offering insights into underlying regulatory mechanisms and biological pathways. However, the sparsity, noise, and the spatial dependency of spatial transcriptomics data pose great analytical challenges. A growing number of computational methods have been developed for constructing spatial gene co-expression networks from spatial transcriptomics (ST) data, including SpaceX [11], Smoothie [12], and hdWGCNA [13]. Unlike traditional co-expression approaches, these models are specifically designed to construct tissue context-specific co-expression networks across cellular and spatial hierarchies, allowing for the preservation of both molecular interactions and spatial organization. By capturing spatially informed co-expression patterns, these methods enable more biologically meaningful representations of gene networks, which can be further leveraged for downstream analyses such as module detection, spatial domain identification, and association with phenotypic traits. Choosing appropriate methods to extract spatial co-expression networks from high-resolution spatial transcriptomics data is crucial for extracting deep biological information, especially as the field shifts toward high-resolution platforms and large-scale studies.

Predicting patient-level clinical outcomes from spatial omics data is an emerging direction for precision medicine, driven by the growing availability of spatially resolved cohorts with linked clinical annotations. Recent spatial-omics-based studies have begun to connect tissue-level spatial patterns with patient phenotypes, including interpretable patient-level prediction from spatial transcriptomics, phenotype-driven discovery using knowledge-guided spatial features, and clinical phenotype modeling from spatial tissue profiles [14–16]. These approaches highlight the potential of spatial omics for population-scale prediction. However, many of them emphasize spatial domains, image-like tissue representations, learned spatial embeddings, or selected molecular signatures, rather than using the spatially informed molecular network features. In addition, many predictive pipelines have limited interpretability to reveal quantitative contributions of essential genes or protein markers, and enriched biological pathways related to clinical outcome.

To address this gap, we propose SPIN (Spatial Predictive Integration Network), a modular framework for population-scale analysis of spatial omics networks and clinical phenotypes. SPIN is designed around three complementary modules: (i) construction of subject-specific spatial co-expression or co-abundance networks; (ii) molecular-to-clinical prediction using BSNMani, a Bayesian scalar-on-network regression model with manifold learning [17]; and (iii) visualization and biological interpretation of phenotype-associated subnetworks, key genes or protein markers, and enriched pathways. Within SPIN, BSNMani currently serves as the predictive modeling engine by decomposing each subject-level network into shared latent subnetworks and subject-specific loadings, which are then associated with clinical outcomes while adjusting for covariates. We present real data examples demonstrating this capability on the MERFISH and clinical data from the Seattle Alzheimer’s Disease Brain Cell Atlas (SEA-AD) study [18,19], and the Imaging Mass Cytometry (IMC) and clinical data from Jackson et al.’s Breast Cancer study [20]. Across these applications, SPIN achieves robust predictive performance while simultaneously revealing meaningful and interpretable biological processes related to the diseases under study.

## Materials and Methods

### Spatial co-expression network generation in SPIN

To characterize transcriptomic coordination among genes, we constructed gene co-expression networks using four approaches: a conventional weighted gene co-expression network analysis (WGCNA) [21] as a baseline method, as well as SpaceX [11], Smoothie [12], and hdWGCNA [13]. WGCNA was applied to normalized gene expression matrices without incorporating spatial coordinates, capturing global transcriptome-wide correlation patterns based on Pearson correlation coefficients and hierarchical clustering of gene modules. Smoothie takes spatial transcriptomics data and outputs gene co-expression modules by smoothing expression, computing spatial correlations, and clustering significant gene pairs. hdWGCNA uses pseudobulk gene expression aggregated by clusters or spatial domains and outputs co-expression modules using the WGCNA framework. SpaceX uses spatial gene expression and coordinates to infer sparse gene co-expression networks through a spatial Poisson model.

### Description of BSNMani method in SPIN

The core of SPIN is BSNMani, a novel Bayesian scalar-on-network regression model designed to jointly analyze high-dimensional networks and clinical scalar outcomes [17]. It decomposes each subject’s high-dimensional networks (e.g., co-expression network) into a low-rank representation and integrates such representation into a regression framework to predict clinical outcomes, while adjusting for any additional clinical covariates. This unified modeling framework facilitates both biological interpretation and accurate prediction of clinical variables. BSNMani comprises two main components, the network model and the clinical regression model. In the first component, BSNMani decomposes each subject’s high-dimensional networks *Y_i_* ∈ R*^N×N^* into a weighted sum of population-shared subnetworks. Specifically, each network matrix is modeled as:

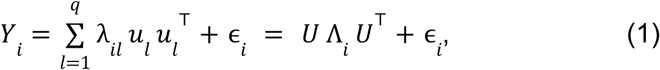

where *U*=[*u_1_*,…,*u_q_*]∈R*^N×q^* is an orthonormal matrix, where each column captures the basis for latent subnetworks shared across subjects, and Λ_i_=diag(λ_i_) is a diagonal matrix encoding subject-specific subnetwork contributions as summarizing network features. The residual matrix *ɛ_i_* is a symmetric matrix capturing individual-level noise with i.i.d. element-wise normal distribution with variance σ^2^ on the off-diagonal. Notably, the orthogonality constraint U^⊤^U=I_q_ ensures that the columns of *U* form an orthonormal frame, and hence lie on the Stiefel manifold *V_q,N_*. This manifold structure enables BSNMani to capture the intrinsic geometric relationships among the latent subnetworks.

The second component models the scalar clinical outcome *C_i_* ∈ R as a linear function of the subject’s network representation λ_i_ and additional clinical covariates *z_i_*∈ R^r^:

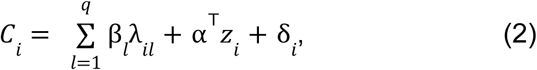

where β = (β1, …, βq)ᵀ ∈ R_q_ and α∈R_r_ are regression coefficients linking network features and covariates to the outcome, and *δ_i_*∼*N*(*0*,*τ^2^*) is the residual error. This regression framework allows the model to predict the clinical phenotype directly using network features, while adjusting for potentially confounding clinical variables. Specifically, by projecting the high-dimensional spatial co-expression networks onto the shared Stiefel manifold, BSNMani summarizes the spatial topology into patient-specific subnetwork loadings, λ*_i_*. These loadings serve as predictive covariates in Equation (2). Consequently, the regression coefficients β*_l_* measure the association between the *l*-th latent spatial subnetwork and the clinical outcome, providing a clear connection between localized spatial patterns and patient-level phenotypes.

BSNMani assigns a uniform prior on the Stiefel manifold to the subnetwork basis parameter *U,* and conjugate priors for the remainder of the model parameters. For posterior inference, BSNMani employs a novel hybrid MALA-Gibbs algorithm [17], using MALA for *U* since it has no closed analytical form and Gibbs algorithm for the remaining parameters. To better facilitate sampling on the Stiefel manifold *V_q,N_*, BSNMani applies one-to-one polar expansion on *U*, projecting it to an *N x q* full rank matrix *X* on the unconstrained Euclidean manifold and circumventing orthogonality constraints:

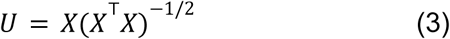

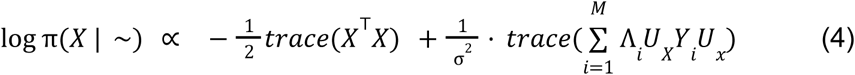

In addition to joint Gibbs-MALA sampling, BSNMani also provides a novel two-stage sampling strategy to further facilitate convergence in high-dimensional cases. In the first stage, BSNMani samples subnetwork basis U, network features λ_i_, and the noise variance σ^2^ using only the observed networks {Y_i_}. In the second stage, BSNMani treats the posterior samples of λ_i_ from the first stage as fixed covariates and infers only the clinical regression parameters via Gibbs sampling. Finally, BSNMani employs an additional Metropolis-Hastings step incorporating the updated normalizing constant based on λ to approximate joint sampling [17].

### Seattle Alzheimer’s Disease Brain Cell Atlas (SEA-AD) MERFISH dataset

We utilized spatial transcriptomics data generated by multiplexed error-robust fluorescence in situ hybridization (MERFISH) from the SEA-AD project [18]. For spatial transcriptomic profiling, MERFISH was applied to the middle temporal gyrus (MTG) of 27 SEA-AD donors meeting the selection criteria of having matched MERFISH expression data, spatial coordinates, clinical metadata, and cell-type annotations. resulting in 69 total brain sections [19]. The SEA-AD MERFISH panel profiled more than 300,000 segmented cells at near-single-cell resolution. Cells were assigned to molecularly defined subclasses based on transcriptomic signatures, consistent with annotations derived from matched single-nucleus RNA-seq (snRNA-seq) datasets. All spatial transcriptomics data and donor-level metadata are publicly available through the Allen Brain Map portal (https://portal.brain-map.org/explore/seattle-alzheimers-diseaase) and the ABC Atlas platform.

The raw MERFISH gene count tables contained 180 feature labels, including 140 panel genes and 40 blank control barcodes. Feature names were screened for mitochondrial-gene, blank-control, and negative-probe patterns; consistent with the SEA-AD MERFISH design, no mitochondrial genes or negative-control probes were detected, and the 40 blank control barcodes were removed before downstream analysis. Detected transcripts not assigned to a segmented cell were discarded, and the remaining records were summed per gene within each cell into a gene-by-cell matrix. Gene–cell entries with no detected transcript were encoded as 0 (a true non-detection, not a missing value to impute [22]). After preprocessing, the resulting expression matrices had an overall sparsity of 65.87%, consistent with targeted single-cell spatial transcriptomics. Cells with fewer than 20 detected genes were excluded using Giotto’s cell-level filtering function before downstream network construction. Spatial coordinates (x, y) for each cell were computed by averaging detected transcript positions. A Giotto object was constructed for each sample to encapsulate raw expression and spatial metadata [22]. Normalization and scaling were performed using total-count normalization (scale factor = 10,000), and both raw and normalized expression matrices were stored for downstream analysis. Cell-type annotation was performed by the SEA-AD consortium, which provides a three-level hierarchy—class (3 types), subclass (24 types), and supertype (138 types). All preprocessing steps were implemented in R using the Giotto, dplyr, and data.table packages within a high-performance computing environment. The resulting gene expression matrices, spatial coordinates, and clinical metadata were serialized as RDS objects for reproducible access.

Clinical annotations were retrieved from the SEA-AD donor metadata file [19] and filtered to include only donors with matched MERFISH profiling of the middle temporal gyrus (MTG). All categorical or ordinal neuropathological traits were numerically encoded for modeling purposes. A binary outcome variable indicating clinical diagnosis of dementia (yes = 1, no = 0) was derived from the SEA-AD consensus clinical diagnostic fields and was used as the primary dependent variable in subsequent logistic regression analyses. To ensure the meaningfulness of the clinical covariates under the constraint of the small sample size, each of them was associated with AD outcome individually, and only those with a significant association were kept for BSNMani modeling (in this case, atherosclerosis).

For the analysis in the main Results, one representative MERFISH section per donor was selected as the section with the largest number of high-quality segmented cells, so that each donor contributed a single subject-specific co-expression network for donor-level model selection and cross-validation. To evaluate robustness to section selection, we additionally performed a sensitivity analysis using all sections of each donor (varying from 1 to 4 sections) that passed preprocessing and network-construction requirements. In this sensitivity analysis, a co-expression network was first constructed separately from each slide of the same donor, then averaged to produce a single donor-level network for BSNMani modeling. This design preserved donor-level analysis while incorporating within-donor section information and preventing information leakage. The per-donor section composition is reported in **Supplementary Table 4**. The all-section sensitivity analysis results are reported in **Supplementary Table 5**, showing the same best q value selection as in the single-section analysis.

### Breast Cancer (BC) Imaging Mass Cytometry (IMC) dataset

We used a single-cell spatial proteomics dataset derived from the IMC platform in a previous study on Breast Cancer pathology [20]. To ensure sufficient cellular resolution, we removed samples with low cell counts, defined as those below the first quartile (Q1) of the log-transformed distribution (log1p < 7.4, ∼1,635 cells). The number of filtered segmented cells per patient ranges from 1,647 to 6,909. Additionally, patients with missing age or survival information (e.g., dead/alive status, overall survival time in months) were excluded. After quality control and data cleaning, data from 253 patients remained, with associated clinical covariates including ER, PR, HER2 status, age, tumor grade, and pathological stage. The cohort composition and clinical characteristics are summarized in **Supplementary Figure 3**.

Each patient sample includes single-cell measurements of 50 metal-tagged proteins at single-cell resolution within spatial context. Unlike spatial transcriptomics, spatial proteomics platforms measure continuous metal-conjugated antibody intensities rather than discrete transcript counts, so exact zeros are rare. Our evaluation of the IMC breast cancer dataset exhibited 0% sparsity and contained no missing values, thus imputation was unnecessary.

The metal tag-protein marker-gene mapping links each measured metal isotope tag to its corresponding protein marker and the associated gene, enabling integration of proteomic measurements with gene-level biological information. Background metal tags and proteins with near-zero variance across cells were excluded, resulting in a refined panel of 29 informative protein markers spanning immune, epithelial, and other functional categories. Intensity matrices were z-score normalized across cells for each protein marker to mitigate batch effects and enable comparison across patients. Finally, a spatial coordinate matrix of (x, y) positions for each cell was constructed, retaining only cells with matching expression and spatial data.

### Benchmarking and model evaluation

To evaluate model-design choices, we performed comparisons among the spatial network-generation methods, over the various number of latent subnetworks *q*. For network construction, we compared WGCNA, SpaceX, Smoothie, and hdWGCNA for the SEA-AD MERFISH dataset, and WGCNA, Smoothie, and hdWGCNA for the IMC breast cancer dataset; SpaceX was not applied to the IMC dataset due to its computational time in this larger cohort. For BSNMani, candidate q values were selected according to the analysis setting: q ∈ {2,…,5} for the small SEA-AD MERFISH dataset, and q ∈ {2,…,8} for the larger IMC breast cancer dataset. For the oligodendrocyte-specific SEA-AD analysis, hdWGCNA was fixed as the network-construction, as it was the best co-expression construction method in the prior all-cell-type SEA-AD analysis, and only the number of BSNMani latent subnetworks was fine tuned.

Candidate network-generation methods and q values were evaluated under a sample-level cross-validation framework, in which candidate-configuration evaluation, model fitting, held-out prediction, and final performance aggregation were kept separate (**Supplementary Figure 2**). For each configuration and each cross-validation split, the model was fitted using only the training sample and evaluated on the held-out samples or the held-out fold. Final performance for each configuration was calculated by aggregating out-of-fold predictions (for the smaller SEA-AD dataset) or across all folds (for the larger IMC breast cancer dataset). Classification performance on the SEA-AD dataset was evaluated using Accuracy, AUROC, AUPRC, MCC, F1 score, Macro F1, Precision, Recall, and Specificity. Survival prediction in the IMC breast cancer dataset was evaluated using concordance index (C-index) and log-ranked p-values among risk groups. To quantify uncertainty, we performed nonparametric bootstrapping with B = 3,000 resamples over the aggregated out-of-fold prediction pairs. BSNMani is a Bayesian scalar-on-network regression model rather than a neural network and therefore does not require GPU-based training. The hardware environment, software dependencies, and empirical runtimes for spatial network construction and BSNMani inference are detailed in **Supplementary Note 4**.

Additionally, to assess the contribution of BSNMani’s manifold-based scalar-on-network modeling, we compared it with two classical feature-selection penalized regression methods: Lasso and Elastic Net. For BSNMani, subject-specific networks were decomposed into latent subnetwork loadings λᵢ, which were used as predictors together with clinical covariates. For Lasso and Elastic Net, the lower-triangular entries of each subject-specific network matrix were vectorized and concatenated with the same clinical covariates, representing the conventional flattened-feature approaches.

### Identification of subnetwork gene co-expression modules

To study the structure of subnetworks and ensure consistent gene ordering across the heatmaps, we applied the Similarity Network Fusion (SNF) technique to integrate the subnetwork matrices into a single fused matrix [23]. This approach preserves the distinct features of each individual subnetwork in the same order, while enabling effective pattern comparisons. Gaussian Mixture Modeling (GMM) was then performed on the fused matrix to cluster the gene features into expression modules based on similarity in their co-expression profiles [24]. The resulting clustering was used to unify the gene ordering across all subnetwork heatmaps, thereby enhancing interpretability and visual comparison.

### Gene Set Enrichment Analysis (GSEA)

Gene Set Enrichment Analysis (GSEA) was performed on gene co-expression modules detected from each subnetwork to identify biologically relevant pathways. We utilized the enrichR [25] R package to conduct enrichment analysis against KEGG [26] and Gene Ontology (GO) [27] biological process databases. Pathways were filtered based on statistical significance, retaining those with adjusted p-values below 0.05. Enriched pathways were further examined for biological relevance and used to interpret the functional roles of each subnetwork.

### Hub gene and hub protein marker identification

For each BSNMani-derived latent subnetwork, hub genes (SEA-AD) or hub protein markers (IMC) were identified based on the total connectivity within the subnetwork-specific co-expression or co-abundance matrix. The total connectivity (hub score) of each gene or marker was computed as the sum of its edge weights to all other genes or markers in that subnetwork, and features were ranked by this score; the top 15 genes (SEA-AD) or top 10 markers (IMC) were reported as hubs. For hub-centered network visualization, gene–gene pairs with pairwise correlations above the 95th percentile (SEA-AD) or marker–marker correlations above the median (IMC) within each subnetwork were retained.

### Survival Analysis

Survival analysis was conducted using Cox proportional hazards regression [28] to evaluate the association between SPIN-derived spatial proteomic subnetwork loadings and patient survival outcomes. The model included normalized subnetwork loadings as predictors, adjusting for relevant clinical covariates such as age, tumor grade, and clinical subtypes. Risk scores were calculated as linear combinations of estimated coefficients and predictor values. Patients were stratified into high-, intermediate-, and low-risk groups based on tertiles of the risk score. Kaplan-Meier curves were generated to visualize survival differences between groups, and statistical significance was assessed using the log-rank test. Model performance was evaluated by the concordance index (C-index), with risk-score directionality explicitly scaled so that higher risk scores corresponded to earlier event times.

## Results

### Overview of SPIN framework

SPIN is a modular framework for population-scale clinical prediction and biological discovery from spatial omics networks (**Figure 1**). Instead of analyzing spatial transcriptomic or proteomic profiles solely as within-sample measurements, SPIN represents each subject’s spatial omics profile as a subject-specific symmetric network and links these networks to clinical phenotypes. The framework consists of three coordinated modules: (i) construction of subject-specific spatial co-expression or co-abundance networks, including gene–gene co-expression networks from MERFISH data and marker–marker co-abundance networks from IMC data; (ii) molecular-to-clinical prediction using BSNMani [17], which models high-dimensional network structures while adjusting for clinical covariates; and (iii) visualization and biological interpretation of phenotype-associated subnetworks, subject-specific loading factors, hub genes or protein markers, and enriched pathways. This modular design enables SPIN to incorporate diverse spatial network-generation methods and clinical endpoints while maintaining a direct connection between patient-level predictions and interpretable spatial molecular patterns.

**Figure 1:**
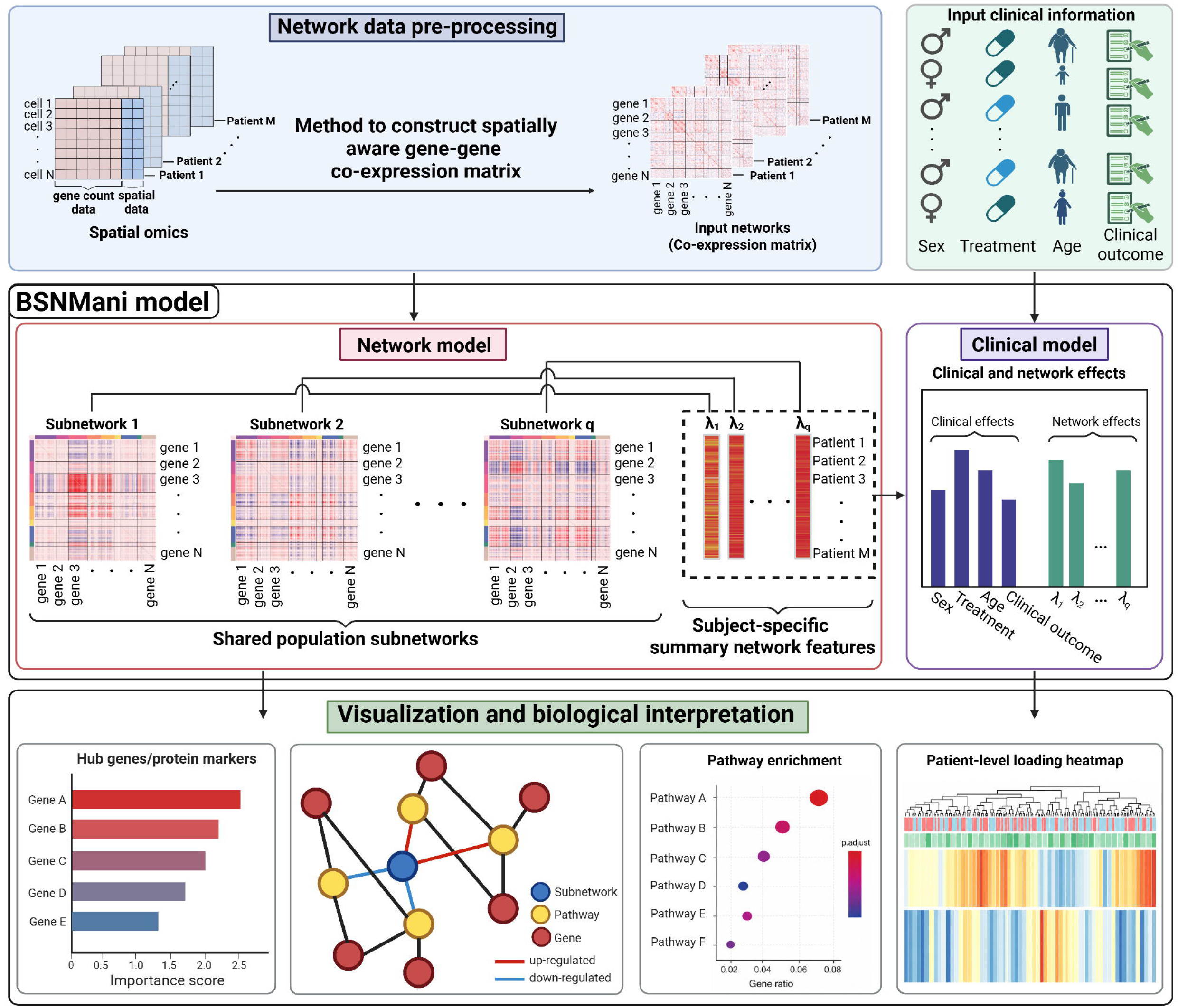
Workflow of SPIN framework. SPIN consists of three modules: i) network construction, ii) BSNMani-based molecular-to-clinical prediction, and iii) visualization/biological interpretation. Spatial omics data are first preprocessed and converted into subject-specific symmetric networks, such as gene–gene co-expression or marker–marker co-abundance matrices. These networks, together with clinical information, are then modeled by BSNMani to estimate shared population subnetworks, subject-specific loading factors, and clinical/network effects. The visualization module summarizes model outputs through hub gene/protein marker ranking plots, subnetwork–gene/marker–pathway networks, gene set enrichment analysis, and heatmaps on clinical variables and loading factors among patients.

Within SPIN, BSNMani serves as the molecular-to-clinical prediction module. It decomposes each input network **Yᵢ** ∈ ℝ^NˣN^ into a linear combination of *q* shared population-level subnetworks. These subnetworks, encoded in the orthonormal basis matrix **U** ∈ ℝ^Nˣq^, capture latent functional structures that are common across individuals. Subject-specific subnetwork loadings **λᵢ** ∈ ℝ^q^ quantify the extent to which each latent subnetwork contributes to the individual’s observed network. This decomposition not only reduces dimensionality but also yields biologically interpretable features that summarize network variation at the subject level. In the second component, the estimated subnetwork loadings λᵢ are used as predictors in a regression model to estimate their associations with clinical outcomes, while adjusting for clinical covariates such as sex, age, and treatment. This scalar-on-network regression framework enables the identification of subnetworks that are predictive of the clinical phenotype, providing insights into the molecular mechanisms underlying disease heterogeneity. Overall, BSNMani offers a unified and scalable approach for integrating high-dimensional symmetric network data and clinical phenotypes, while maintaining biological interpretability.

### Benchmarking on gene co-expression network method and feature selection method on SEA-AD MERFISH dataset

In spatial omics research, constructing accurate gene co-expression networks is essential for elucidating spatial regulatory mechanisms. BSNMani is a regression model that incorporates sample-specific co-expression networks as key inputs. The performance of this model depends on how these co-expression matrices are constructed. While conventional methods such as WGCNA have been widely used to construct co-expression networks in non-spatial transcriptomics data, they do not incorporate spatial proximity between cells and thus may be suboptimal for spatially resolved single-cell data. We thus compared several space-aware co-expression network construction methods, including SpaceX [11], Smoothie [12], and hdWGCNA [13] against the baseline method WGCNA, on SEA-AD MERFISH data.

A close examination of the co-expression matrices reveals that Smoothie and hdWGCNA produce clearer block structure than WGCNA, whereas SpaceX produces weaker or less stable co-expression patterns in the representative donor-level matrix (**Figure 2a**). Additional donor-level heatmap comparisons across all four network-construction methods are shown in **Supplementary Figure 4**. Moreover, for each of the four co-expression matrix construction methods (WGCNA, hdWGCNA, Smoothie, SpaceX), we varied the number of subnetworks *q* among the population in BSNMani decomposition (*q* = 2, 3, 4, or 5) on SEA-AD MERFISH data. The objective is to predict dementia status among SEA-AD donors, using logistic regression as the second modeling step of BSNMani. Given the small sample size (n = 27), we employed leave-one-out cross-validation (LOOCV) to assess model performance. Nine metrics—Accuracy, Precision, Recall, F1 score, Specificity, Area Under the Receiver Operating Characteristic (AUROC), Area Under the Precision-Recall Curve (AUPRC), Matthews Correlation Coefficient (MCC), and macro F1 scores—were used to comprehensively evaluate the predictive power of each *q* configuration.

**Figure 2:**
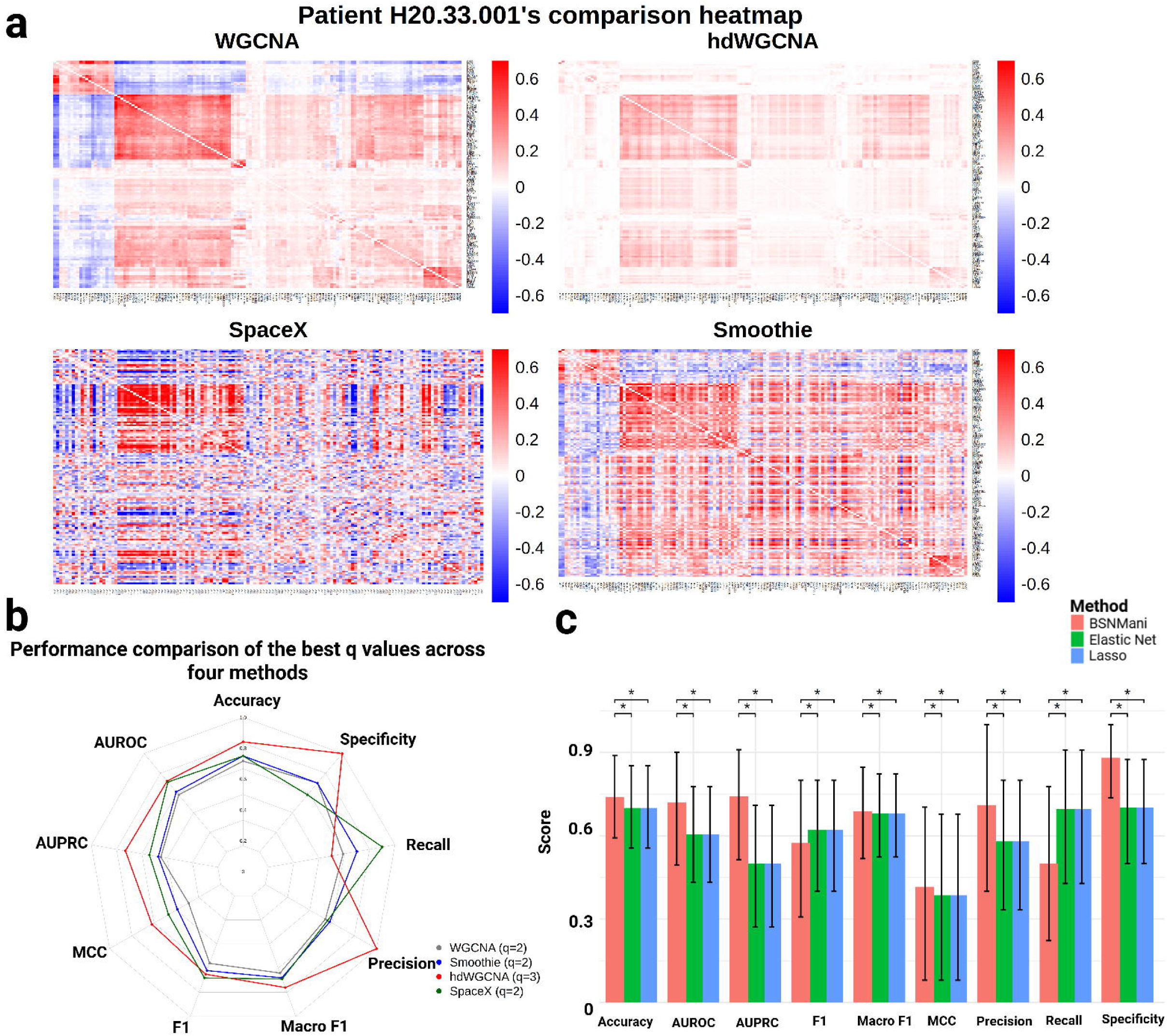
Selection of the best spatial co-expression generation methods using the SEA-AD MERFISH data. a) Gene co-expression matrices, for example, patient H20.33.001 constructed using different spatial co-expression construction methods: WGCNA, hdWGCNA, SpaceX, and Smoothie. b) Performance comparison of the best q values across four methods. c) Benchmark results on feature selection methods using BSNMani, Elastic Net and Lasso.

Among all combinations of *q* values and network construction methods, hdWGCNA with *q* = 3 yields the best overall performance, achieving an accuracy of 0.81, an MCC score of 0.62, and an AUPRC score of 0.73 **(Figure 2b, Supplementary Table 1)**. It is better than other network-construction methods with various *q* values. Notably, co-expression matrices obtained from WGCNA, the non-spatial baseline method, only achieve an accuracy of 0.66 and an AUPRC of 0.45 in the BSNMani model when *q* = 2. Thus, incorporating spatial context leads to a clear improvement in predicting dementia status among SEA-AD donors. Based on these results, we selected hdWGCNA with *q* = 3 as the final SEA-AD configuration for downstream prediction and biological interpretation.

For clinically relevant gene feature selection, we compared BSNMani with two other methods: Lasso and Elastic Net for their performance. As shown in **Figure 2c**, under the same LOOCV framework, BSNMani achieves higher Accuracy, AUROC, AUPRC, MCC, macro F1 score, Precision, and Specificity than both Lasso and Elastic Net (bootstrap-based Wilcoxon tests, p < 0.05). In particular, the mean Precision and AUROC for BSNMani are 0.71 and 0.72 respectively, as compared to 0.58 and 0.58 for Lasso and 0.60 and 0.60 for Elastic Net. These results support the methodological advantage of subnetwork-based representation of spatial co-expression data in BSNMani over classical penalized regression models.

### Interpretation of SPIN model on AD prediction using MERFISH SEA-AD data

BSNMani yields gene-gene co-expression subnetworks and their corresponding loading vector λᵢ, which is used to construct the clinical model to predict donor phenotype. We thus focus on their interpretations, exemplified by the best AD prediction BSNMani model (*q* = 3) using MERFISH SEA-AD data. Each subnetwork shows distinct co-expression structures, enriched biological functions, and network hub-gene profiles (**Figure 3a–c**), allowing direct tracing of biological functions related to dementia-status prediction.

**Figure 3:**
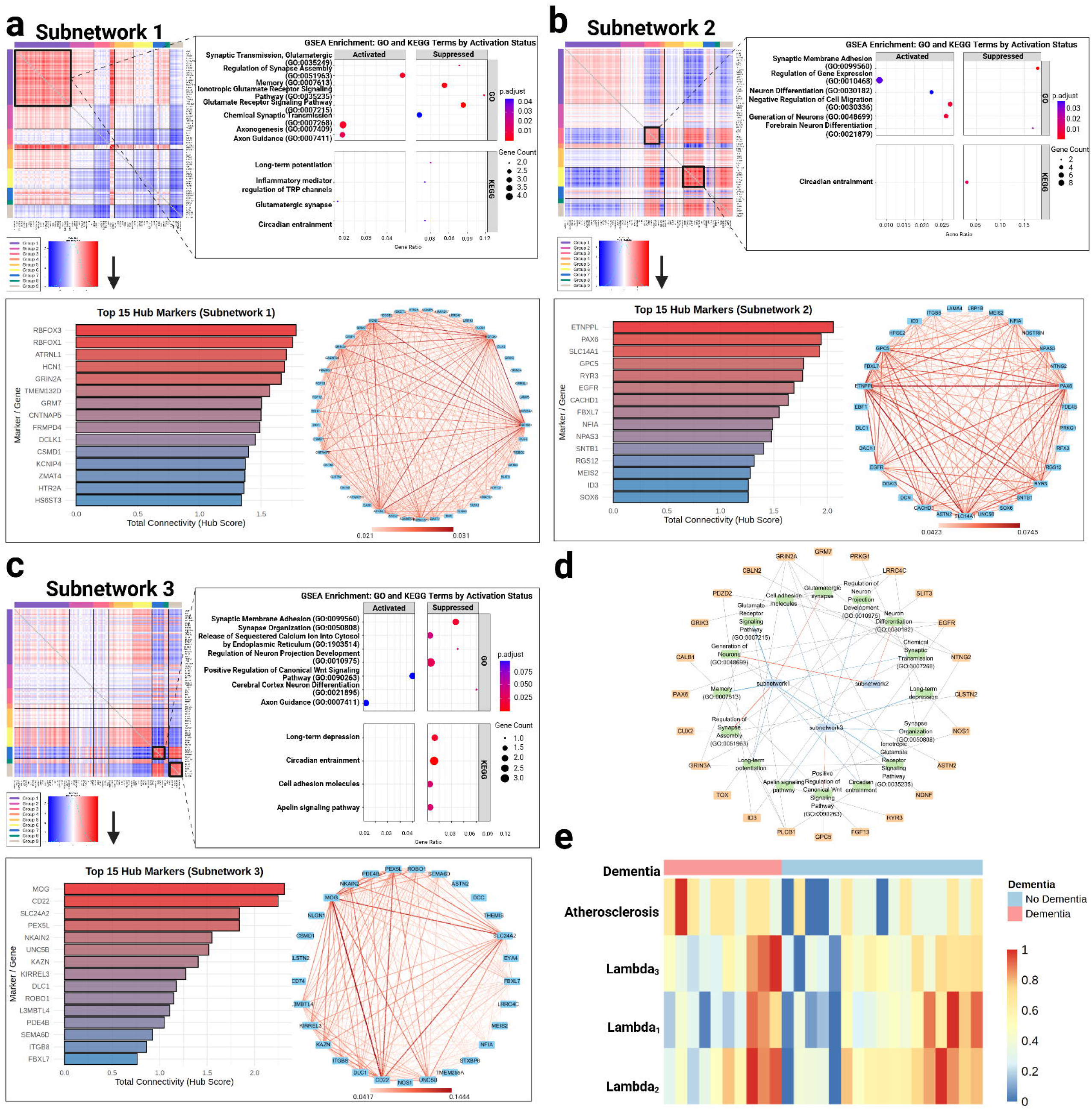
Application of SPIN to the MERFISH SEA-AD dataset. a-c) Subnetwork-specific visualization for the three optimal BSNMani-derived latent co-expression subnetworks (q = 3), each including the co-expression heatmap, pathway enrichment bubble plot, hub-gene ranking bar plot, and gene–gene correlation network visualization based on the top 95th percentile of pairwise correlations. d) Network visualization of key genes and their connections across subnetworks (red edge: activated; blue edge: suppressed). e) Heatmap of values on atherosclerosis and the three lambda values (after Min-Max normalization) in the final SPIN framework. Donors are grouped by dementia status. The color bar on the left indicates dementia status (red: Dementia, blue: non-Dementia).

Subnetwork 1 shows clear co-expression modules and is associated with neuronal and synaptic processes, such as synaptic transmission and assembly, axonogenesis, long-term potentiation, inflammatory mediator regulation of TRP channels, and glutamate receptor signaling (**Figure 3a**). Hub-gene analysis further identifies highly connected genes in the subnetwork, including RBFOX3, RBFOX1, ATRNL1, HCN1, and GRIN2A, supporting the neuronal and synaptic relevance of this subnetwork. Subnetwork 2 captures a distinct co-expression pattern and is enriched for pathways related to synaptic membrane adhesion, regulation of gene expression and cell migration, and neuron differentiation (**Figure 3b**). The top hub genes include ETNPPL, PAX6, SLC14A1, GPC5, and RYR3. On the other hand, subnetwork 3 shows additional co-expression modules enriched for release of sequestered calcium ion into cytosol by endoplasmic reticulum, regulation of neuron projection development, Wnt signaling pathway, and apelin signaling pathway (**Figure 3c**). The top hub genes include MOG, CD22, SLC24A2, PEX5L, and NKAIN2.

We further generated an integrated network visualization linking BSNMani-derived subnetworks, hub genes, and enriched pathways (**Figure 3d**). Such visualization summarizes how the latent network features used for dementia prediction connect to specific genes and pathway-level biological processes. Finally, the clinical and subnetwork-loading heatmap shows that the final logistic regression model includes four predictive variables: atherosclerosis and the three BSNMani-derived subnetwork loadings, λ_1_, λ_2_, and λ_3_ (**Figure 3e** and **Supplementary Note 1**). As expected, atherosclerosis is positively associated with AD outcome, with a positive coefficient of 2.6070. The SPIN-derived subnetwork loadings also contribute to the final model, with a positive coefficient for λ3 (β3 = 0.6651), but negative coefficients for λ1 and λ2 (β1 = −0.3519 and β2 = −0.0941). Consistent with this model, donors with higher λ3 values show greater association with dementia status (**Figure 3e)**. Together, these results show that SPIN not only predicts dementia status from spatial transcriptomic networks but also provides interpretable and meaningful information on subnetworks, hub genes, and enriched biological pathways associated with AD.

### Patient survival prediction by SPIN using a breast cancer cohort with Imaging Mass Cytometry (IMC) data

To evaluate the generalizability of SPIN across single-cell modalities and disease contexts, we applied it to predict survival in 253 breast cancer patients whose tumor tissues underwent single-cell IMC profiling with 29 protein markers. Among the evaluated network-construction methods and subnetwork numbers (q values), Smoothie with q = 2 achieves the best survival prediction performance, with a bootstrapped mean C-index of 0.78 (95% CI: 0.66–0.87) and log-rank p = 1.09 × 10⁻⁹ for the three stratified risk groups (**Figure 4a, b** and **Supplementary Table 2**). In comparison, the Cox-PH model with clinical-data only (age, tumor grade, and clinical subtype) achieves a C-index of 0.708 and log-rank p = 1.56 × 10⁻⁸ (**Figure 4c**), confirming that the SPIN-derived subnetwork loadings provide prognostic information beyond the clinical covariates.

**Figure 4:**
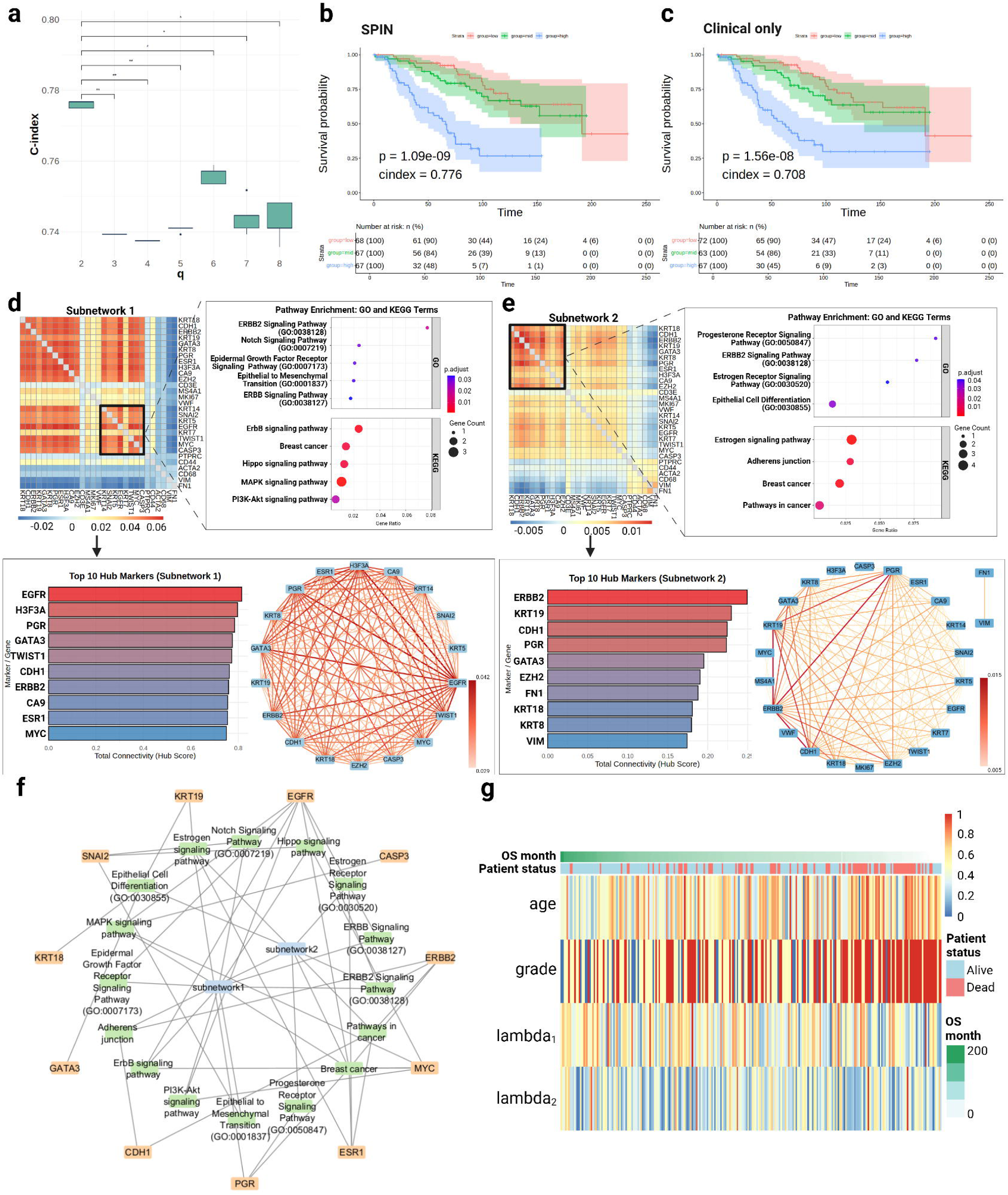
Application of SPIN to IMC breast cancer spatial proteomics cohort. a) Model predictive performance (C-index) across latent subnetworks (*q* ∈ {2, ···, 8}) using Smoothie based network construction and BSNMani pipeline. b-c) Kaplan-Meier survival curves for patients stratified into risk groups (Low, Intermediate, High), using the survival prediction from (b) SPIN framework and (c) the clinical-variable-only Cox-PH model. Metrics shown are C-index (cindex) and log-rank p-values of the KM curves among risk-groups for each model. d-e) Subnetwork-specific visualization for the two SPIN-derived latent subnetworks, including marker–marker co-abundance heatmaps (boxed region: module markers used for pathway enrichment), pathway enrichment bubble plots, top 10 hub-marker ranking plots, and marker–marker correlation network visualization based on correlations above the median threshold. f) Integrated network visualization of representative protein markers, their enriched pathways, and their connections across the two subnetworks (gray edges link markers to pathways and pathways to subnetworks). g) Heatmap of clinical covariates (age, grade) and λ-subnetwork covariates across patients, ordered by overall survival time (OS months, longest to shortest) and annotated with survival status (alive vs. dead).

We further analyzed the biological functions of the two SPIN-derived subnetworks based on pathway enrichment of their module markers, hub-marker connectivity, and their associations with overall survival (**Figures 4d–e**). The two subnetworks correspond to overlap and distinct molecular programs of breast cancer. Subnetwork 1 is enriched for various signaling pathways (eg. ErbB/EGFR, MAPK, PI3k-Akt, Hippo and Notch), and is characterized by hub markers EGFR, PGR, GATA3, ERBB2, ESR1 (**Figure 4d**). This suggests a proliferative state of the subnetwork, consistent with the more favorable clinical behavior of hormone-receptor-positive, luminal-like breast cancers. Such “relatively protective” effect is further supported by the negative Cox-PH coefficient for λ₁ (β₁ = −0.5837) and HR = 0.56 (**Supplementary Note 2**). On the other hand, subnetwork 2, representing a luminal-epithelial hormone-receptor program plus EMT/mesenchymal program, presents an overall weak adverse association with survival (β₂ = 0.0612, HR = 1.06). The hub proteins include the luminal-epithelial markers ERBB2, KRT19, KRT18, KRT8, PGR, and GATA3, E-cadherin (CDH1), as well as aggressive phenotype markers including mesenchymal markers FN1 and VIM, and the polycomb regulator EZH2 (**Figure 4e**). The unique biological events of this subnetwork include progesterone signaling pathways, epithelial cell differentiation and CDH1-mediated adherens junctions. The tripartite network graph in **Figure 4f** integrates the markers, enriched pathways, and subnetworks, and confirms the shared as well as unique programs between the two subnetworks. Lastly, the heatmap shows the λ predictors and clinical covariates along patients’ overall survival time (**Figure 4g**). Consistent with the signs of the coefficients shown earlier, patients with higher λ₁ values exhibit longer overall survival, confirming the protecting effect, whereas higher λ₂ values are associated with shorter survival durations and higher mortality.

### Interpretation of SPIN model on AD prediction using cell-type specific MERFISH SEA-AD data

Cell–type specific level analysis provides additional granularity for linking spatial co-expression networks with patient-level phenotypes. We demonstrate this capability using SEA-AD MERFISH data, as they include well-annotated cell types. Oligodendrocyte is the most abundant cell type in the SEA-AD MERFISH cohort **(Supplementary Figure 1)**, and it has been repeatedly implicated in Alzheimer’s disease pathogenesis, warranting prioritization for cell–type–specific analysis [29]. Recent work shows that oligodendrocytes themselves produce amyloid-β and contribute to plaque burden in AD models, further underscoring their direct involvement in AD pathology [30]. Thus, we constructed oligodendrocyte-specific co-expression networks for SPIN modeling, with the same objective of predicting dementia status among SEA-AD donors. We chose hdWGCNA for co-expression network construction, the best-performing method in the main SEA-AD all-cell analysis (**Figure 2b**). All downstream SPIN modeling and interpretation steps followed the same workflow as the previous SEA-AD analysis of all cell types in **Figure 3**. Among the different q values for the number of subnetworks, the configuration of q = 2 achieves the best overall performance balance under LOOCV, with an accuracy of 0.70, MCC of 0.39, F1 score of 0.62, and macro F1 score of 0.68 **(Supplementary Table 3)**. This result shows that oligodendrocyte-specific spatial co-expression networks retain clinically relevant information despite arising from a subset of cells.

The two oligodendrocyte-specific subnetworks reveal very distinct co-expression patterns and biological functions. Subnetwork 1 shows striking repression on multiple biological presses, including synapsis transmission, glutamate receptor signaling, calcium signaling/transmembrane transportation (**Figure 5a**). Hub-gene analysis identifies highly connected genes including RBFOX1/3, HS3ST2, HCN1, and CLSTN2, confirming that this subnetwork captures oligodendrocyte-associated signals related to synaptic regulation, calcium signaling, and neuron–glia communication. On the other hand, subnetwork 2 has the opposite activation trends on synapse organization/assembly and cell adhesion (**Figure 5b**). The top hub genes include MOG, CD22, UNC5B, PDE4B, and DLC1, highlighting oligodendrocyte-associated molecular features related to myelination and cell adhesion. We further generated an integrated network visualization connecting the oligodendrocyte-specific subnetworks, hub genes, and enriched pathways (**Figure 5c**). Moreover, the donor-level heatmap of BSNMani-derived loading factors shows Atherosclerosis and λ1 are positively associated with dementia status with β values of 1.6582 and 0.1328, respectively, whereas λ2 has a negative association with dementia status with its β value of −0.2749 (**Figure 5d, Supplementary Note 3**). Together, these results demonstrate that SPIN can be applied at the cell-type-specific level to recover biologically interpretable subnetworks, providing finer resolution of disease-associated signals across cell types.

**Figure 5:**
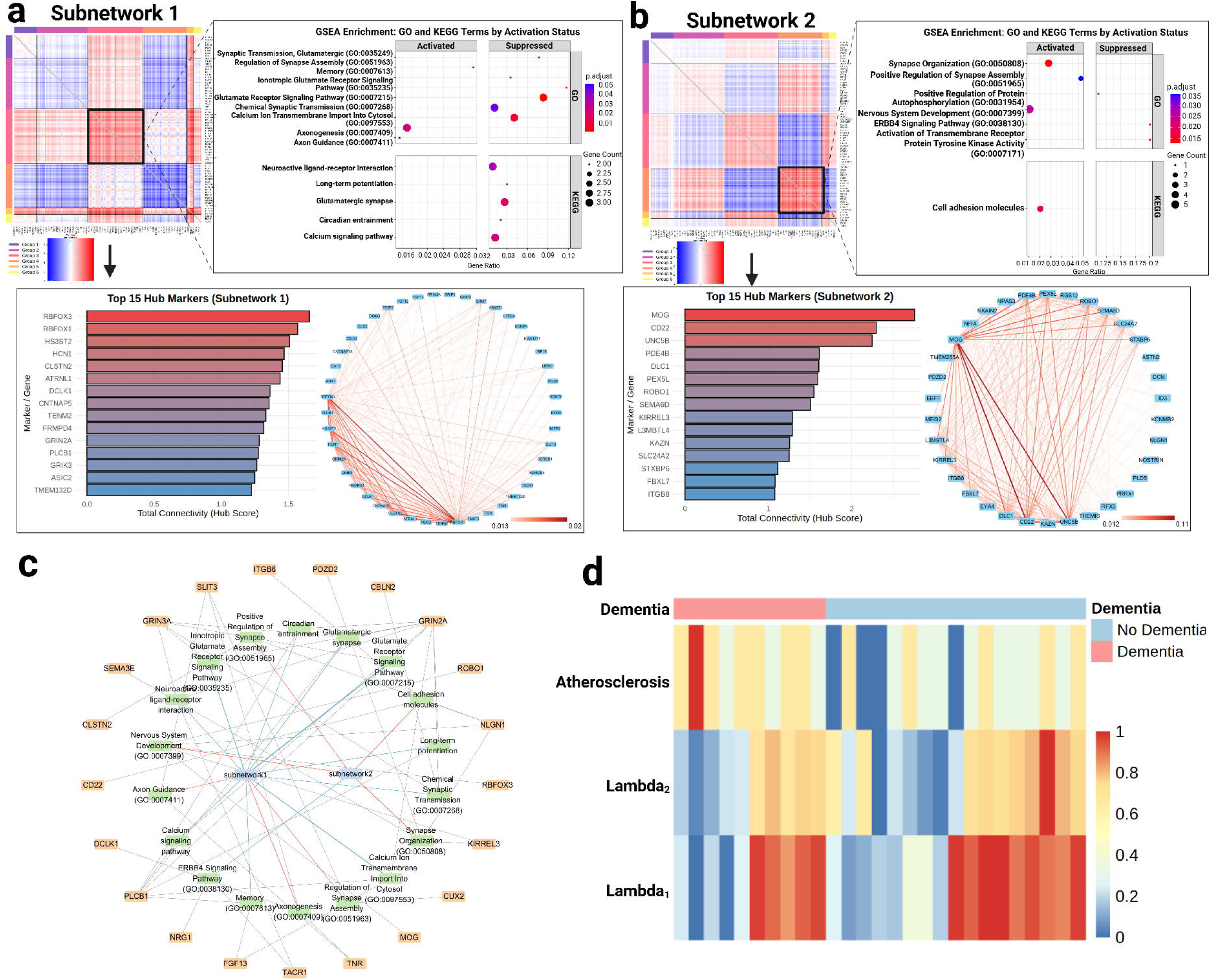
Application of SPIN to oligodendrocyte cell-type-specific SEA-AD MERFISH data. a-b) Subnetwork-specific visualization for the two BSNMani-derived latent co-expression subnetworks (q = 2), including co-expression heatmaps, pathway enrichment bubble plots, hub-gene ranking plots, and gene–gene correlation network visualizations based on the top 95th percentile of pairwise correlations. c) Network visualization of key gene hubs and their connections across subnetworks (red edge: activated; blue edge: suppressed). d) Heatmap integrating clinical covariates and λ-subnetwork covariates, with individuals organized by dementia versus non-dementia status.

## Discussion

In this study, we developed SPIN as a modular population-scale framework for linking spatial omics-derived molecular networks with clinical phenotypes. SPIN represents each subject by a spatially informed co-expression or co-abundance network, models population-level network variation using BSNMani, and connects the resulting latent subnetworks with clinical outcomes while retaining a direct path to biological interpretation. We demonstrate this framework across two single-cell-resolution modalities, MERFISH spatial transcriptomics and IMC spatial proteomics, showing its applicability to both dementia-status classification and breast cancer survival prediction.

In the network construction module, we showed that methods including hdWGCNA and Smoothie are more desirable for constructing spatially aware gene co-expression networks at the population level. hdWGCNA incorporates spatial coordinates into network construction by aggregating neighboring cells into spatially coherent metacells and computing spatially weighted gene–gene correlations. On the other hand, Smoothie leverages spatial neighborhoods to apply kernel smoothing to the expression matrix, thereby amplifying stable covariation within homogeneous microdomains and suppressing technical noise, which improves the consistency and signal-to-noise ratio of edge-weight estimates. It captures microenvironment-driven co-regulation patterns and reduces technical noise, yielding more biologically grounded features that improve the downstream predictive performance. The choice of hdWGCNA vs. Smoothie is likely dependent on factors such as data type (e.g., spatial transcriptomics data, spatial proteomics data, etc), cohort size, and study objective (e.g., classification, survival prediction). It will be of interest for future studies to compare more systematically.

A central component of SPIN is BSNMani, which represents each subject-specific network through population-shared latent subnetworks and subject-specific loading factors [17]. In the SEA-AD benchmark, BSNMani outperforms Lasso and Elastic Net when all methods were evaluated using the same network inputs, clinical covariates, and subject-level cross-validation framework. Lasso and Elastic Net use vectorized network edges as individual predictors, whereas BSNMani first decomposes each subject-specific network into a small set of shared subnetworks among subpopulations and uses the resulting subject-specific loadings (λ) as predictors. This subnetwork-level representation, estimated under Stiefel-manifold constraints with a hybrid MALA/Gibbs strategy, concentrates signal into biologically coherent modules and mitigates variance inflation from highly correlated edges, thereby improving AUROC, accuracy, precision, specificity, and several summary performance metrics.

The biological findings from the SEA-AD analysis were consistent with major processes implicated in Alzheimer’s disease and dementia. Despite the modest cohort size of 27 donors, SPIN reached an accuracy of 0.81 for dementia-status prediction, and revealed three subnetworks collectively captured alterations in neuronal remodeling and axonal program [31], synaptic and glutamatergic signaling [32–34], neurogenesis and synaptic-adhesion regulation [35,36], calcium homeostasis and synaptic integrity [37,38], Wnt-related neuroprotection [39], and circadian regulation [40]. These themes align with known neurodegenerative mechanisms involving impaired synaptic function and neuronal communication together with compensatory remodeling responses. SPIN can also be applied at the cell-type-specific level. In the oligodendrocyte-specific SEA-AD analysis, SPIN preserved clinically relevant predictive information while revealing subnetworks associated with neuron–glia communication, calcium-dependent oligodendrocyte function, myelination-related cell adhesion, and trophic ERBB4 signaling [33]. These patterns were diluted in the all-cell analysis, illustrating how cell-type-specific network construction provides complementary resolution into disease-associated molecular programs (**Supplementary Note 5, Supplementary References S1–S5**).

The IMC breast cancer analysis similarly identified spatial proteomic subnetworks associated with established aspects of tumor biology and prognosis. The top markers and enriched processes reflected hormone-receptor and ERBB/EGFR growth-factor signaling [41,42], epithelial differentiation and epithelial-to-mesenchymal transition with Hippo-related regulation [43,44], and E-cadherin-mediated cell adhesion [45]. These findings suggest that the patient-level survival signal captured by SPIN reflects coordinated variation across tumor-intrinsic, epithelial-state, and immune-associated programs rather than individual protein markers alone. In short, across two studies, SPIN connects biological programs through the subject-specific subnetwork loadings with clinical prediction, providing an interpretable molecular link between population-level network variation and clinical phenotypes.

Spatial omics data can be sparse and noisy, particularly at single-cell resolution. SPIN mitigates this at two levels. First, quality-control filtering is applied before network construction, removing low-quality cells (e.g., those with very few detected genes) so that unreliable measurements do not propagate into the networks. Second, rather than modeling isolated, cell-level counts, SPIN summarizes each subject’s spatial omics profile into a subject-specific co-expression network. This macroscopic, network-level representation reduces reliance on individual sparse gene–cell entries and emphasizes stable, population-level structure. Moreover, spatially aware network construction methods, such as hdWGCNA and Smoothie, leverage information from neighboring cells through metacell aggregation or kernel smoothing across spatial neighborhoods. By stabilizing edge-weight estimates and mitigating localized dropout, these approaches enable inferred subnetworks to better represent underlying biological signals rather than technical noise.

While SPIN provides a robust framework for population-scale spatial omics, several considerations remain for future development. First, the framework depends on the quality and resolution of the input spatial omics data and the reliability of the subject-specific networks constructed from them. Extremely low-resolution platforms, exceptionally high cellular sparsity, or weak spatial organization may reduce the stability of inferred networks and therefore downstream prediction. Second, although SPIN is designed to be platform-agnostic, broader validation across independent cohorts, additional disease contexts, and emerging spatial technologies (e.g., 10x Visium HD or Slide-seq) will be important to establish generalizability beyond the two datasets evaluated here,consistent with recent cross-dataset and cross-platform evaluation studies in spatial transcriptomics [46,47]. Future studies with larger cohorts and more extensive within-subject spatial sampling will enable more systematic evaluation of regional heterogeneity.

The modular design of SPIN also provides opportunities for methodological expansion. Because network construction, molecular-to-clinical prediction, and biological interpretation are separable components, complementary approaches can be incorporated at different stages of the workflow. For example, spatially variable gene selection methods such as SpatialDE can be included as an upstream module for filtering spatial gene features [4]. Other emerging population-level spatial feature-phenotype modeling approaches [14–16] can be incorporated into SPIN as well, when their input representations and prediction objectives are compatible. Additionally, new spatial multi-omics technologies, including DBiT-seq-based spatial transcriptome-protein co-profiling [48] and spatial epigenome-transcriptome co-profiling [49], may further enable extension of SPIN with integrated modeling in the future. Thus, SPIN is intended as an open and inclusive framework that can evolve with advances in spatial network inference and clinical prediction methodology.

In summary, SPIN provides a modular and interpretable strategy for integrating subject-specific spatial molecular networks with clinical phenotypes at the population level. Across SEA-AD MERFISH and breast cancer IMC data, SPIN supported clinical prediction while identifying biologically interpretable subnetworks associated with disease-relevant molecular programs, providing a foundation for broader validation and future extension of population-scale spatial omics network modeling.

## Author’s contributions

L.X.G. conceived this project, supervised the study and participated in the manuscript writing. Y.L. co-supervised the study, assisted with analysis and participated in the manuscript writing. T.L., Y.Y., H.W., and E.K. carried out the analysis. T.L and E.K. wrote the main manuscript, with the participation of Y.Y and H.W. T.U. assisted with the analysis, interpretation and participated in the manuscript writing. S.W. and JK assisted with the analysis.

## Acknowledgements

The authors thank all lab members of Garmire Group for helpful discussions.

## Data availability

Raw data for the SEA-AD dataset are available at https://registry.opendata.aws/allen-sea-ad-atlas/, and raw data for the IMC Breast Cancer dataset are available at https://zenodo.org/records/3518284. Processed and derived data generated in this study, including co-expression matrices generated by different methods and BSNMani model outputs, are deposited in Zenodo under https://zenodo.org/records/17400565. All code used to produce the results of this study is available at GitHub: https://github.com/lanagarmire/SPIN.

