## Supplementary Materials for "Population-Scale Integration of Spatial Omics Networks for Clinical Prediction and Biological Discovery by SPIN"

### Supplementary Figure 1: Cell-type proportions in the SEA-AD MERFISH dataset.

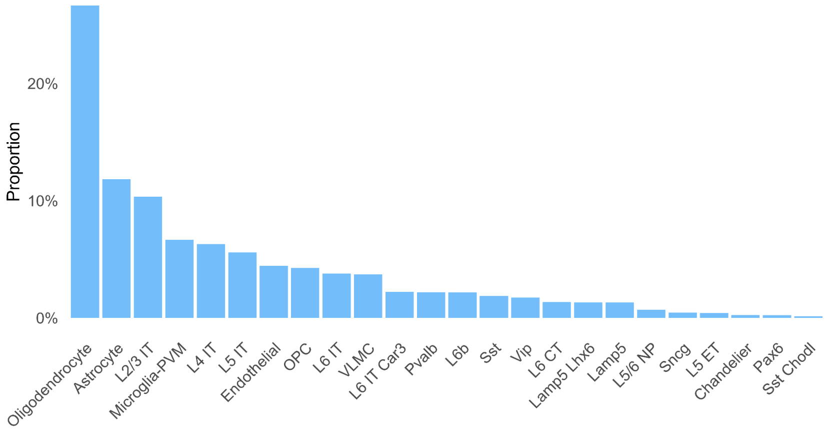


### Supplementary Figure 2: Workflow for SPIN-based population-level prediction from spatial omics networks.

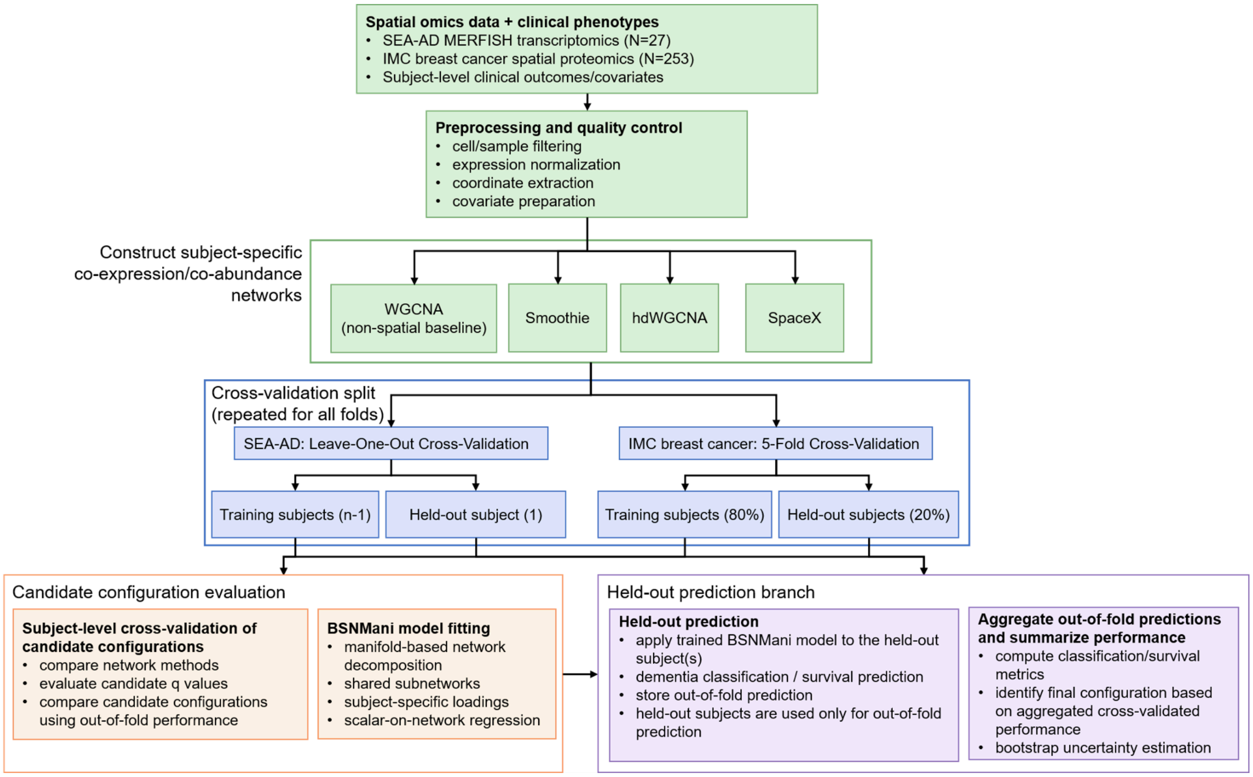

**Supplementary Figure 3: Summary of the IMC breast cancer spatial proteomics cohort.**

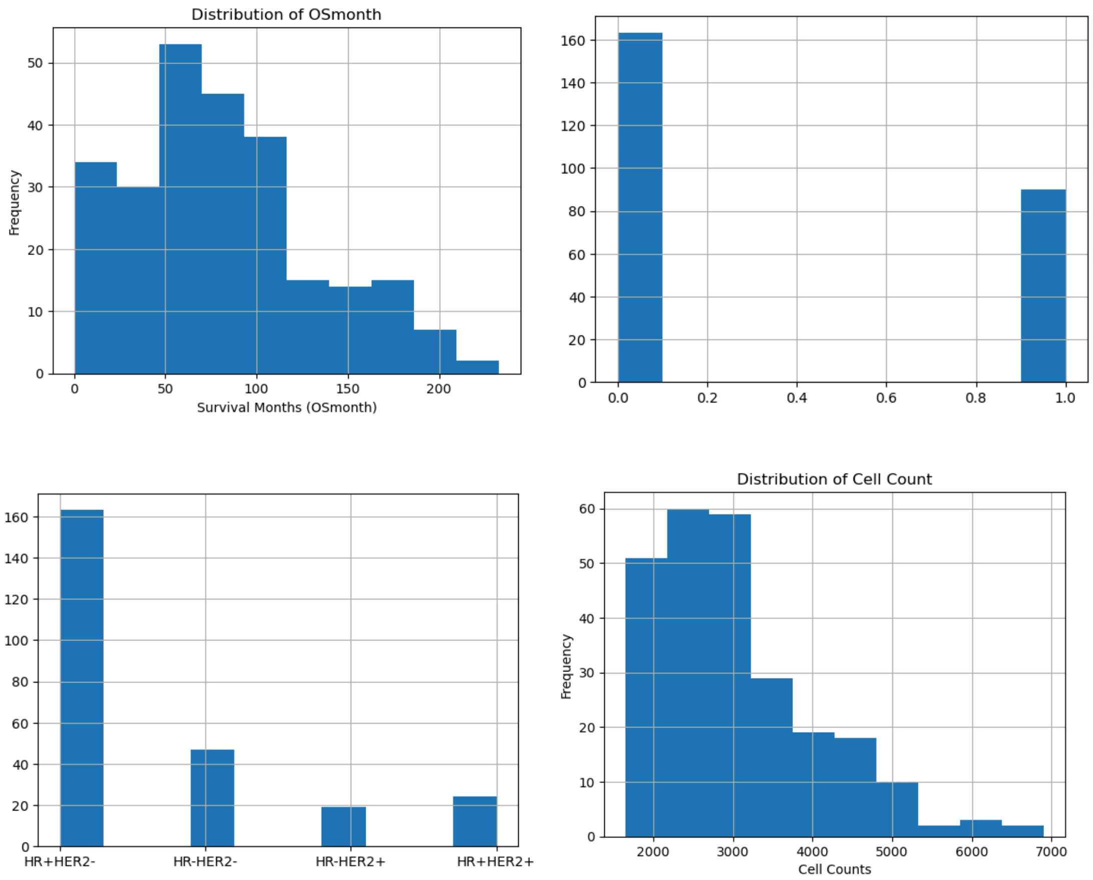

**Supplementary Figure 4: Additional donor-level co-expression heatmap comparisons across four network-construction methods in SEA-AD MERFISH data.**

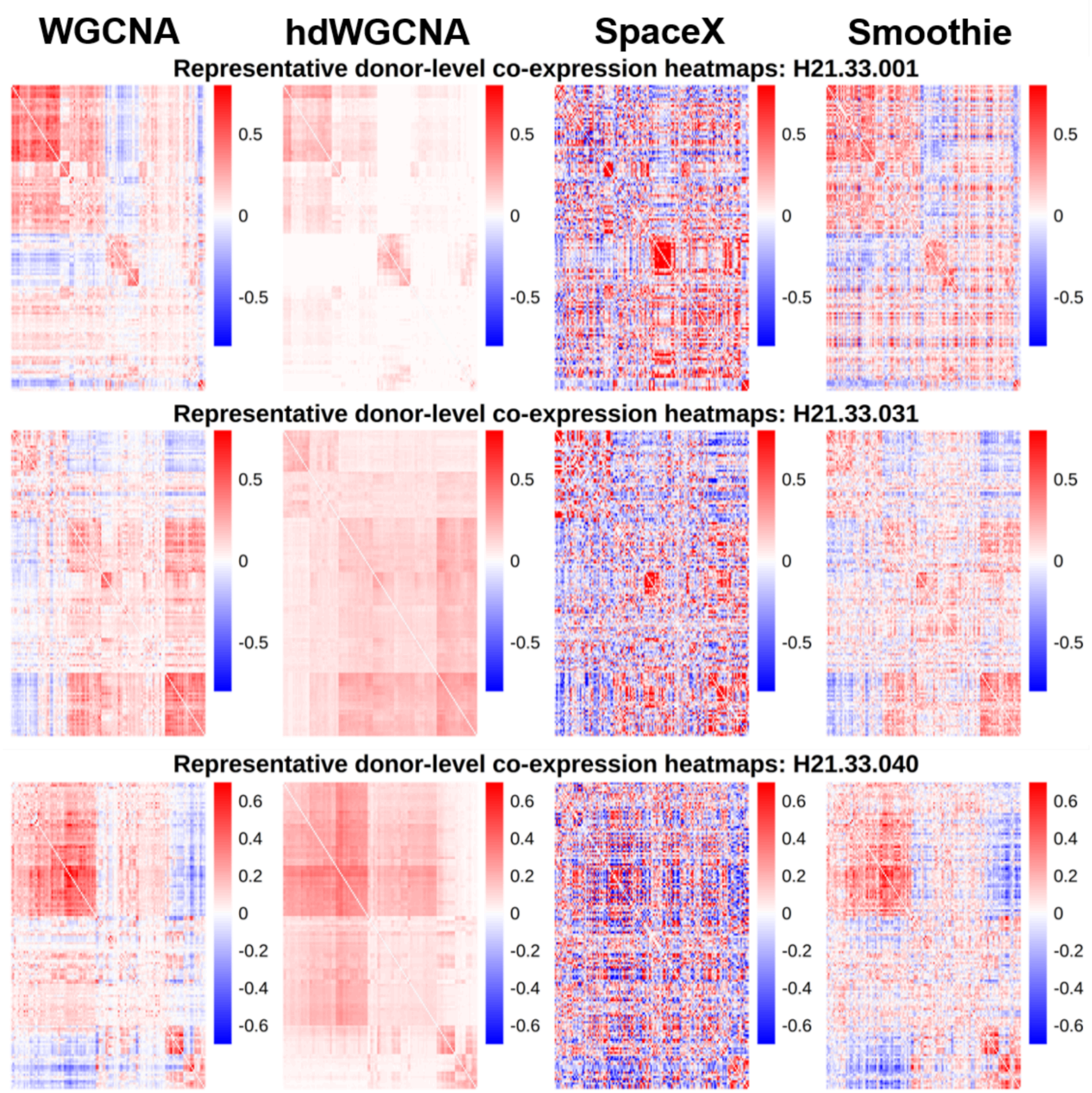

**Supplementary Table 1: Selection of spatial co-expression network construction methods for the SEA-AD MERFISH dataset.**

Nine evaluation metrics—Accuracy, Area Under the Receiver Operating Characteristic Curve (AUROC), Area Under the Precision–Recall Curve (AUPRC), Matthews Correlation Coefficient (MCC), F1-score, Macro F1-score, Precision, Recall, and Specificity—were used to comprehensively assess the predictive performance of each q configuration (q = 2–5) using leave-one-out cross-validation (LOOCV). Reported results represent the mean values and 95% confidence intervals derived from 3,000 bootstrap resamples.

| Method | $q$ | Accuracy | AUROC | AUPRC | MCC | F1 Score | Macro F1 | Precision | Recall | Specificity |
|---|---|---|---|---|---|---|---|---|---|---|
| WGCNA | 2 | 0.66 (0.48, 0.81) | 0.58 (0.39, 0.78) | 0.45 (0.19, 0.71) | 0.29 (-0.10, 0.64) | 0.56 (0.29, 0.79) | 0.64 (0.44, 0.81) | 0.54 (0.25, 0.83) | 0.59 (0.28, 0.90) | 0.70 (0.47, 0.90) |
| | 3 | 0.63 (0.44, 0.81) | 0.61 (0.43, 0.81) | 0.44 (0.20, 0.73) | 0.32 (-0.03, 0.63) | 0.60 (0.36, 0.80) | 0.62 (0.44, 0.80) | 0.50 (0.25, 0.73) | 0.80 (0.50, 1.00) | 0.53 (0.29, 0.75) |
| | 4 | 0.63 (0.44, 0.81) | 0.62 (0.46, 0.82) | 0.42 (0.20, 0.71) | 0.32 (-0.03, 0.64) | 0.61 (0.36, 0.81) | 0.62 (0.44, 0.81) | 0.50 (0.25, 0.75) | 0.80 (0.50, 1.00) | 0.53 (0.29, 0.77) |
| | 5 | 0.63 (0.44, 0.81) | 0.56 (0.36, 0.79) | 0.34 (0.16, 0.55) | 0.23 (-0.15, 0.59) | 0.53 (0.25, 0.76) | 0.60 (0.42, 0.78) | 0.50 (0.21, 0.77) | 0.59 (0.29, 0.90) | 0.64 (0.40, 0.87) |
| Smoothie | 2 | 0.70 (0.52, 0.85) | 0.61 (0.42, 0.82) | 0.47 (0.20, 0.74) | 0.39 (0.02, 0.71) | 0.62 (0.33, 0.83) | 0.68 (0.49, 0.85) | 0.58 (0.29, 0.85) | 0.70 (0.38, 1.00) | 0.70 (0.47, 0.90) |
| | 3 | 0.63 (0.44, 0.81) | 0.58 (0.41, 0.78) | 0.45 (0.18, 0.71) | 0.23 (-0.16, 0.60) | 0.53 (0.25, 0.77) | 0.60 (0.41, 0.79) | 0.50 (0.20, 0.78) | 0.60 (0.27, 0.90) | 0.64 (0.40, 0.87) |
| | 4 | 0.67 (0.48, 0.85) | 0.64 (0.42, 0.85) | 0.53 (0.24, 0.81) | 0.30 (-0.06, 0.65) | 0.56 (0.29, 0.80) | 0.64 (0.46, 0.81) | 0.54 (0.25, 0.83) | 0.60 (0.29, 0.90) | 0.70 (0.47, 0.92) |
| | 5 | 0.63 (0.44, 0.81) | 0.60 (0.43, 0.70) | 0.41 (0.19, 0.72) | 0.23 (-0.17, 0.60) | 0.53 (0.25, 0.77) | 0.61 (0.42, 0.80) | 0.50 (0.20, 0.78) | 0.60 (0.27, 0.90) | 0.64 (0.40, 0.87) |
| hdWGCNA | 2 | 0.70 (0.52, 0.85) | 0.61 (0.42, 0.82) | 0.52 (0.22, 0.80) | 0.33 (-0.06, 0.68) | 0.54 (0.22, 0.78) | 0.65 (0.46, 0.84) | 0.62 (0.25, 1.00) | 0.49 (0.17, 0.80) | 0.82 (0.63, 1.00) |
| | 3 | 0.81 (0.67, 0.96) | 0.72 (0.44, 0.94) | 0.73 (0.45, 0.93) | 0.62 (0.37, 0.87) | 0.65 (0.31, 0.91) | 0.76 (0.56, 0.93) | 1.00 (1.00, 1.00) | 0.50 (0.16, 0.83) | 1.00 (1.00, 1.00) |
| | 4 | 0.59 (0.41, 0.78) | 0.58 (0.42, 0.76) | 0.44 (0.18, 0.71) | 0.17 (-0.20, 0.53) | 0.51 (0.24, 0.73) | 0.57 (0.39, 0.76) | 0.46 (0.18, 0.73) | 0.59 (0.29, 0.90) | 0.58 (0.33, 0.81) |
| | 5 | 0.78 (0.59, 0.93) | 0.65 (0.45, 0.88) | 0.56 (0.25, 0.88) | 0.51 (0.13, 0.82) | 0.61 (0.29, 0.88) | 0.72 (0.53, 0.91) | 0.83 (0.43, 1.00) | 0.50 (0.18, 0.83) | 0.94 (0.80, 1.00) |
| SpaceX | 2 | 0.70 (0.52, 0.85) | 0.71 (0.50, 0.90) | 0.54 (0.26, 0.86) | 0.47 (0.17, 0.74) | 0.68 (0.44, 0.86) | 0.69 (0.51, 0.85) | 0.56 (0.31, 0.80) | 0.90 (0.67, 1.00) | 0.58 (0.35, 0.81) |
| | 3 | 0.63 (0.44, 0.81) | 0.62 (0.43, 0.83) | 0.46 (0.20, 0.76) | 0.32 (-0.04, 0.64) | 0.61 (0.35, 0.80) | 0.62 (0.43, 0.81) | 0.50 (0.25, 0.75) | 0.80 (0.50, 1.00) | 0.53 (0.29, 0.77) |
| | 4 | 0.66 (0.48, 0.81) | 0.60 (0.42, 0.81) | 0.43 (0.19, 0.73) | 0.33 (-0.04, 0.65) | 0.59 (0.33, 0.80) | 0.65 (0.46, 0.81) | 0.54 (0.25, 0.80) | 0.70 (0.38, 1.00) | 0.64 (0.40, 0.87) |
| | 5 | 0.56 (0.37, 0.74) | 0.60 (0.44, 0.79) | 0.40 (0.19, 0.68) | 0.36 (0.18, 0.55) | 0.62 (0.40, 0.79) | 0.53 (0.36, 0.72) | 0.45 (0.25, 0.65) | 1.00 (1.00, 1.00) | 0.29 (0.07, 0.52) |

**Supplementary Table 2: Selection of spatial co-abundance network construction methods for the IMC breast cancer dataset.**

| Method | $q$ | Bootstrapped Mean C-index (95% CI) | P-value |
|---|---|---|---|
| WGCNA | 2 | 0.71 (0.59, 0.82) | $9.53 \times 10^{-7}$ |
| | 3 | 0.73 (0.59, 0.85) | $1.46 \times 10^{-7}$ |
| | 4 | 0.73 (0.59, 0.85) | $1.28 \times 10^{-9}$ |
| | 5 | 0.72 (0.58, 0.85) | $2.78 \times 10^{-9}$ |
| | 6 | 0.70 (0.55, 0.82) | $6.36 \times 10^{-9}$ |
| | 7 | 0.69 (0.54, 0.82) | $6.89 \times 10^{-9}$ |
| | 8 | 0.67 (0.54, 0.80) | $6.21 \times 10^{-11}$ |
| Smoothie | 2 | 0.78 (0.66, 0.87) | $1.09 \times 10^{-9}$ |
| | 3 | 0.74 (0.62, 0.85) | $2.50 \times 10^{-8}$ |
| | 4 | 0.74 (0.61, 0.85) | $9.72 \times 10^{-10}$ |
| | 5 | 0.74 (0.62, 0.85) | $6.51 \times 10^{-10}$ |
| | 6 | 0.76 (0.64, 0.86) | $3.59 \times 10^{-9}$ |
| | 7 | 0.75 (0.63, 0.85) | $4.80 \times 10^{-8}$ |
| | 8 | 0.74 (0.62, 0.85) | $1.62 \times 10^{-7}$ |
| hdWGCNA | 2 | 0.74 (0.60, 0.86) | $3.09 \times 10^{-10}$ |
| | 3 | 0.69 (0.55, 0.82) | $9.37 \times 10^{-8}$ |
| | 4 | 0.71 (0.57, 0.84) | $3.31 \times 10^{-11}$ |
| | 5 | 0.70 (0.57, 0.82) | $2.28 \times 10^{-10}$ |
| | 6 | 0.70 (0.56, 0.82) | $9.78 \times 10^{-10}$ |
| | 7 | 0.68 (0.54, 0.80) | $1.69 \times 10^{-9}$ |
| | 8 | 0.72 (0.58, 0.84) | $3.04 \times 10^{-9}$ |

**Supplementary Table 3: Selection of q in the oligodendrocyte-specific SEA-AD analysis using hdWGCNA-derived co-expression networks.** hdWGCNA was selected as the best-performing network-construction method, based on previous analysis on all cell types in SEA-AD data. Candidate q values from 2 to 5 were evaluated using donor-level leave-one-out cross-validation.

| Method | $q$ | Accuracy | AUROC | AUPRC | MCC | F1 Score | Macro F1 | Precision | Recall | Specificity |
|---|---|---|---|---|---|---|---|---|---|---|
| hdWGCNA | 2 | 0.70 (0.52, 0.85) | 0.59 (0.39, 0.80) | 0.43 (0.19, 0.71) | 0.39 (0.02, 0.71) | 0.62 (0.33, 0.83) | 0.68 (0.49, 0.85) | 0.58 (0.29, 0.85) | 0.70 (0.38, 1.00) | 0.70 (0.47, 0.90) |
| | 3 | 0.63 (0.44, 0.81) | 0.58 (0.40, 0.77) | 0.43 (0.17, 0.69) | 0.23 (-0.16, 0.61) | 0.53 (0.25, 0.77) | 0.61 (0.42, 0.80) | 0.50 (0.20, 0.78) | 0.60 (0.27, 0.90) | 0.64 (0.41, 0.86) |
| | 4 | 0.66 (0.48, 0.81) | 0.61 (0.42, 0.82) | 0.45 (0.19, 0.76) | 0.33 (-0.04, 0.65) | 0.59 (0.32, 0.80) | 0.65 (0.32, 0.80) | 0.53 (0.25, 0.80) | 0.70 (0.38, 1.00) | 0.64 (0.40, 0.87) |
| | 5 | 0.70 (0.52, 0.85) | 0.61 (0.44, 0.83) | 0.44 (0.20, 0.75) | 0.36 (-0.01, 0.70) | 0.58 (0.31, 0.82) | 0.67 (0.49, 0.85) | 0.60 (0.29, 0.90) | 0.59 (0.29, 0.90) | 0.76 (0.56, 0.94) |

**Supplementary Table 4: Details on SEA-AD MERFISH data by slide.** The table lists matched donor-section samples included in the all-section sensitivity analysis, including donor ID, slide/specimen ID, total cells, and cognitive status, with a total of 60 matched donor-section slides from 27 SEA-AD donors, with one to four sections per donor.

| Donor ID | Slide ID | Total Cells | Status | Donor ID | Slide ID | Total Cells | Status |
|---|---|---|---|---|---|---|---|
| H20.33.001 | 1194111462 | 54,404 | 0 | H21.33.013 | 1116997297 | 11,606 | 1 |
| H20.33.001 | 1217500585 | 54,556 | 0 | H21.33.014 | 1217501462 | 28,642 | 0 |
| H20.33.001 | 1217500590 | 58,314 | 0 | H21.33.015 | 1162693722 | 22,991 | 0 |
| H20.33.004 | 1185930845 | 25,728 | 1 | H21.33.015 | 1162693727 | 23,072 | 0 |
| H20.33.004 | 1185930850 | 20,611 | 1 | H21.33.015 | 1162693732 | 19,894 | 0 |
| H20.33.004 | 1218552387 | 30,485 | 1 | H21.33.016 | 1217499815 | 17,679 | 1 |
| H20.33.004 | 1218552392 | 35,615 | 1 | H21.33.016 | 1217499820 | 16,835 | 1 |
| H20.33.012 | 1218552479 | 17,380 | 0 | H21.33.016 | 1217499825 | 18,242 | 1 |
| H20.33.012 | 1218552485 | 22,195 | 0 | H21.33.019 | 1238559004 | 16,865 | 0 |
| H20.33.015 | 1296199271 | 24,885 | 1 | H21.33.019 | 1238559483 | 17,478 | 0 |
| H20.33.015 | 1296199276 | 27,886 | 1 | H21.33.021 | 1170797669 | 5,518 | 1 |
| H20.33.015 | 1296199281 | 22,343 | 1 | H21.33.022 | 1182389655 | 9,591 | 0 |
| H20.33.025 | 1176833728 | 15,797 | 0 | H21.33.022 | 1182389662 | 9,896 | 0 |
| H20.33.025 | 1176833739 | 18,613 | 0 | H21.33.022 | 1182389669 | 9,796 | 0 |
| H20.33.025 | 1176833749 | 14,818 | 0 | H21.33.023 | 1218552275 | 13,322 | 0 |
| H20.33.035 | 1194111807 | 44,444 | 0 | H21.33.023 | 1218552293 | 20,092 | 0 |
| H20.33.035 | 1194111812 | 43,736 | 0 | H21.33.023 | 1218552298 | 16,153 | 0 |
| H20.33.040 | 1195928136 | 20,736 | 1 | H21.33.025 | 1218552010 | 25,500 | 0 |
| H20.33.044 | 1172983297 | 16,410 | 0 | H21.33.025 | 1218552030 | 27,138 | 0 |
| H20.33.044 | 1172983307 | 20,801 | 0 | H21.33.028 | 1195927756 | 8,428 | 0 |
| H21.33.001 | 1192523789 | 32,444 | 1 | H21.33.028 | 1217500948 | 7,161 | 0 |
| H21.33.001 | 1192523794 | 33,388 | 1 | H21.33.028 | 1217500958 | 5,852 | 0 |
| H21.33.005 | 1217500871 | 8,973 | 1 | H21.33.031 | 1296510469 | 56,977 | 1 |
| H21.33.005 | 1217500876 | 11,141 | 1 | H21.33.031 | 1296510477 | 33,450 | 1 |
| H21.33.006 | 1175046724 | 4,917 | 0 | H21.33.032 | 1196150174 | 16,082 | 0 |
| H21.33.006 | 1175046730 | 13,600 | 0 | H21.33.038 | 1218552100 | 8,172 | 0 |
| H21.33.011 | 1217500806 | 21,453 | 0 | H21.33.038 | 1218552111 | 11,251 | 0 |
| H21.33.011 | 1217500810 | 20,089 | 0 | H21.33.040 | 1296199298 | 14,227 | 0 |
| H21.33.011 | 1217500814 | 20,871 | 0 | H21.33.040 | 1296199308 | 16,228 | 0 |
| H21.33.012 | 1174511271 | 16,161 | 1 | H21.33.040 | 1296199314 | 22,176 | 0 |

*Note:* Cognitive Status 0 = No Dementia, 1 = Dementia.

**Supplementary Table 5: Performance metrics using all slides in the SEA-AD MERFISH cohort.** The slides are detailed in Supplementary Table 4. Candidate network-construction methods and q values were evaluated using donor-level LOOCV. Reported values represent mean performance and 95% confidence intervals from 3,000 bootstrap resamples.

| Method | $q$ | Accuracy | AUROC | AUPRC | MCC | F1 Score | Macro F1 | Precision | Recall | Specificity |
|---|---|---|---|---|---|---|---|---|---|---|
| WGCNA | 2 | 0.63 (0.44, 0.81) | 0.60 (0.41, 0.80) | 0.49 (0.20, 0.75) | 0.27 (-0.10, 0.62) | 0.57 (0.31, 0.79) | 0.61 (0.43, 0.80) | 0.50 (0.23, 0.77) | 0.70 (0.38, 1.00) | 0.59 (0.33, 0.81) |
| | 3 | 0.66 (0.48, 0.81) | 0.60 (0.42, 0.81) | 0.47 (0.20, 0.73) | 0.33 (-0.04, 0.67) | 0.60 (0.33, 0.81) | 0.65 (0.46, 0.81) | 0.54 (0.25, 0.80) | 0.70 (0.38, 1.00) | 0.64 (0.40, 0.86) |
| | 4 | 0.63 (0.44, 0.81) | 0.62 (0.44, 0.82) | 0.45 (0.20, 0.74) | 0.32 (-0.03, 0.63) | 0.60 (0.35, 0.80) | 0.62 (0.44, 0.80) | 0.50 (0.25, 0.73) | 0.80 (0.50, 1.00) | 0.53 (0.29, 0.76) |
| | 5 | 0.59 (0.41, 0.78) | 0.56 (0.33, 0.81) | 0.34 (0.15, 0.57) | 0.18 (-0.21, 0.54) | 0.51 (0.24, 0.74) | 0.57 (0.39, 0.75) | 0.46 (0.19, 0.73) | 0.60 (0.29, 0.90) | 0.58 (0.33, 0.81) |
| Smoothie | 2 | 0.70 (0.52, 0.85) | 0.62 (0.42, 0.83) | 0.51 (0.21, 0.79) | 0.39 (0.03, 0.71) | 0.62 (0.35, 0.83) | 0.68 (0.50, 0.85) | 0.58 (0.29, 0.85) | 0.70 (0.38, 1.00) | 0.70 (0.47, 0.90) |
| | 3 | 0.67 (0.48, 0.81) | 0.61 (0.42, 0.81) | 0.51 (0.20, 0.78) | 0.33 (-0.04, 0.67) | 0.60 (0.32, 0.81) | 0.65 (0.46, 0.81) | 0.54 (0.25, 0.80) | 0.70 (0.38, 1.00) | 0.64 (0.40, 0.87) |
| | 4 | 0.59 (0.41, 0.78) | 0.61 (0.44, 0.81) | 0.43 (0.20, 0.72) | 0.27 (-0.11, 0.59) | 0.58 (0.33, 0.79) | 0.58 (0.40, 0.78) | 0.47 (0.24, 0.71) | 0.80 (0.50, 1.00) | 0.47 (0.24, 0.71) |
| | 5 | 0.74 (0.56, 0.89) | 0.64 (0.44, 0.87) | 0.53 (0.24, 0.86) | 0.45 (0.09, 0.78) | 0.65 (0.38, 0.87) | 0.72 (0.53, 0.89) | 0.64 (0.33, 0.91) | 0.70 (0.38, 1.00) | 0.76 (0.54, 0.94) |
| hdWGCNA | 2 | 0.66 (0.48, 0.81) | 0.63 (0.44, 0.83) | 0.50 (0.21, 0.77) | 0.33 (-0.04, 0.66) | 0.60 (0.33, 0.81) | 0.65 (0.47, 0.81) | 0.54 (0.25, 0.80) | 0.70 (0.38, 1.00) | 0.64 (0.41, 0.87) |
| | 3 | 0.74 (0.56, 0.89) | 0.76 (0.51, 0.93) | 0.60 (0.28, 0.87) | 0.59 (0.38, 0.80) | 0.73 (0.52, 0.90) | 0.73 (0.55, 0.89) | 0.59 (0.35, 0.82) | 1.00 (1.00, 1.00) | 0.59 (0.33, 0.82) |
| | 4 | 0.74 (0.56, 0.89) | 0.64 (0.44, 0.86) | 0.52 (0.23, 0.83) | 0.45 (0.10, 0.78) | 0.65 (0.38, 0.87) | 0.72 (0.54, 0.89) | 0.64 (0.33, 0.91) | 0.70 (0.40, 1.00) | 0.76 (0.54, 0.94) |
| | 5 | 0.63 (0.44, 0.81) | 0.57 (0.37, 0.79) | 0.36 (0.16, 0.62) | 0.20 (-0.19, 0.57) | 0.49 (0.19, 0.73) | 0.59 (0.40, 0.78) | 0.50 (0.17, 0.82) | 0.50 (0.17, 0.82) | 0.70 (0.47, 0.91) |

### Supplementary Note 1. Final logistic regression model for SEA-AD dataset

$$\log\left(\frac{p}{1-p}\right) = -4.4197 + 2.6070 \cdot Atherosclerosis - 0.3519 \cdot \lambda_1 - 0.0941 \cdot \lambda_2 + 0.6651 \cdot \lambda_3$$

$$where\ p = Pr(Y = 1)\ denotes\ the\ probability\ of\ Dementia.$$

### Supplementary Note 2. Final CoxPH model for IMC dataset

$$\eta = 0.5202 \cdot Grade_2 + 1.1876 \cdot Grade_3 + 0.0294 \cdot Age - 0.0101 \cdot ClinicalType_{HR+HER2+}$$

$$- 0.8912 \cdot ClinicalType_{HR-HER2+} + 0.2358 \cdot ClinicalType_{HR-HER2-} - 0.5837 \cdot \lambda_1 + 0.0612 \cdot \lambda_2$$

$$where\ h(t) = h_0(t)exp(\eta)$$

### Supplementary Note 3. Final logistic regression model for the oligodendrocyte-specific SEA-AD dataset

$$\log\left(\frac{p}{1-p}\right) = -2.6438 + 1.6582 \cdot Atherosclerosis + 0.1328 \cdot \lambda_1 - 0.2749 \cdot \lambda_2$$

$$where\ p = Pr(Y = 1)\ denotes\ the\ probability\ of\ Dementia.$$

### Supplementary Note 4. Computational Resources and Model Efficiency

#### Hardware and Software Environment

Unlike highly parameterized deep learning architectures, the SPIN pipeline currently uses BSNMani, a Bayesian scalar-on-network regression framework, as the molecular-to-clinical prediction module. Therefore, the pipeline does not require GPU-based deep learning training, prolonged multi-epoch neural network optimization, or specialized deep learning infrastructure. The main computational costs arise from two steps: subject-specific spatial network construction and Bayesian posterior inference in BSNMani.

All computational experiments, network construction procedures, and model evaluations in this study were performed on the Cheaha supercomputing cluster at the University of Alabama at Birmingham (UAB). Computations were run on the cluster's short partition using Intel Xeon Gold 6248R @ 3.00 GHz processors with up to 384 GB of RAM under a Linux environment managed by the SLURM workload manager. The analytical pipeline was implemented using R version 4.4.0.

#### Empirical Runtimes

The computational footprint of SPIN is primarily distributed across two phases: (1) spatial co-expression or co-abundance network construction and (2) Bayesian posterior inference via Markov chain Monte Carlo (MCMC) sampling in BSNMani.

For spatial network construction, the runtime depends on the number of segmented cells, the number of molecular features, and the selected network-construction algorithm. For the SEA-AD MERFISH transcriptomics cohort, the main analysis included 27 donor-level representative sections, with an average of approximately 19,934 analyzed cells per donor. Constructing the subject-specific spatial co-expression matrices using hdWGCNA required approximately 45 minutes in total. For the larger IMC breast cancer spatial proteomics cohort, which included 253 patients with 1,647 to 6,909 segmented cells per patient, constructing subject-specific spatial co-abundance matrices required approximately 2.5 hours in total.

For BSNMani model training, the core algorithm uses a hybrid MALA-Gibbs sampling strategy to estimate the Stiefel-manifold subnetwork bases, subject-specific subnetwork loadings, and clinical association parameters. For the SEA-AD classification analysis, BSNMani model fitting with 100,000 MCMC iterations was completed in approximately two hours. For the IMC breast cancer Cox proportional hazards survival analysis, BSNMani model fitting with 10,000 MCMC iterations was completed in approximately one hour.

These empirical runtimes demonstrate that SPIN provides a computationally accessible framework for population-level spatial omics analysis. Although we executed the analyses on an institutional high-performance computing cluster for reproducibility and batch processing, the pipeline does not require GPU acceleration or deep learning-specific hardware. Instead, SPIN can be deployed on CPU-based computing environments with sufficient memory, with runtime depending primarily on cohort size, cell count, molecular feature dimension, network-construction method, number of latent subnetworks, and MCMC settings.

**Supplementary Note 5. Extended interpretation of oligodendrocyte-specific subnetworks**

SPIN can be applied at the cell-type-specific level to identify the biological subnetworks associated with donor-level phenotypes within a focal cell type. Using oligodendrocyte lineage in the SEA-AD MERFISH cohort as an example, SPIN not only preserved clinically relevant predictive signals but also revealed additional subnetworks whose enrichment pathways were aligned with oligodendrocyte-associated biology. In the oligodendrocyte-specific analysis, Subnetwork 1 revealed suppression of Glutamate Receptor Signaling Pathway (GO:0007215),

Chemical Synaptic Transmission (GO:0007268), Neuroactive ligand-receptor interaction, and Calcium signaling pathway, indicating impaired neuron–glia communication, disrupted neurotransmission, and altered calcium homeostasis in oligodendrocytes during dementia progression [S1,S2]. In particular, downregulation of Calcium Ion Transmembrane Import Into Cytosol (GO:0097553) and Calcium signaling pathway may reflect impaired $Ca^{2+}$-dependent oligodendrocyte function and defective myelin maintenance, both of which have been implicated in AD-associated white matter degeneration [S3]. Meanwhile, activation of Axonogenesis (GO:0007409) and Regulation of Synapse Assembly (GO:0051963) may represent compensatory remodeling responses attempting to preserve axonal integrity and synaptic support under neurodegenerative stress. Subnetwork 2 was characterized by activation of Synapse Organization (GO:0050808), Positive Regulation of Synapse Assembly (GO:0051965), Nervous System Development (GO:0007399), and Cell adhesion molecules, suggesting active oligodendrocyte involvement in synaptic maintenance and neuron–glia interactions [S4]. However, suppression of ERBB4 Signaling Pathway (GO:0038130) and Activation of Transmembrane Receptor Protein Tyrosine Kinase Activity (GO:0007171) may indicate impaired trophic signaling and disrupted oligodendrocyte maturation pathways in AD [S5]. These results show that the cell-type-specific SPIN framework further increases biological interpretability relative to mixed networks constructed from all cells by identifying biological patterns that are significant only within the focal cell type yet diluted in the overall analysis.
